# Trans Fats Consumption and Body Mass Index in Cross Sectional Analysis

**DOI:** 10.1101/2021.01.13.21249701

**Authors:** Beatrice A. Golomb, Arthur Pavlovsky, Hayley J. Koslik

## Abstract

**Background:** Trans fats remain on the market in parts of the world. Emerging evidence suggests that factors beyond calorie balance may affect fat deposition and body mass index (**BMI**). Trans fats are prooxidant, proinflammatory, and have shown adverse metabolic effects and increased fat deposition in animals.

**Objective:** To assess the relation of dietary trans fatty acid consumption (**dTFA**) to BMI in humans.

**Design:** Cross-sectional analysis of baseline data from a broadly-sampling study.

**Setting:** Community-dwelling adults from Southern California.

**Participants:** 1018 adult men and women aged 20-85 without known diabetes, CVD, or cancer, with screening LDL 115-190mg/dL. Women of procreative potential and children were excluded.

**Measurements:** Height, weight, and waist circumference were measured, and BMI calculated (kg/m^2^). The Fred Hutchinson Food Frequency Questionnaire provided data on dTFA (grams/day) and calories consumed. Additional covariates included activity, chocolate consumption frequency, and mood. Regression assessed the relation of BMI (outcome) to dTFA, unadjusted and in models adjusting for age and sex, and adding calories and activity, chocolate consumption frequency, and mood.

**Results:** Higher dTFA was associated with higher BMI and waist circumference. The coefficient was strengthened, not attenuated, with adjustment for calories and activity, and other assessed covariates. In the fully adjusted model, each gram/day of dTFAs was associated with 0.44 higher BMI (SE=0.12;95%CI=0.21,0.67); P<0.001 and 1cm greater waist circumference (SE=0.35;95%CI=0.37,1.7); P=0.003.

**Conclusion:** Findings relating greater dTFA to higher BMI in humans comport with experimental data in animals, extend adverse metabolic associations of dTFA, and buttress evidence that foods’ composition, as well as caloric number, bears on BMI. Findings are cross-sectional but strength and consistency of association, biological gradient, and biological plausibility add “weight” to the prospect of a causal connection.

**Strengths and limitations of this study:**

- Findings are cross-sectional and rely on dietary recall.
- Observational studies carry risks of bias and confounding; but randomized trials are problematic where products with potential for harm are under evaluation.
- Although some nations have implemented plans to remove or reduce dTFAs from the food supply, dTFA remain on the market in other nations, rendering findings of continued interest.
- The association of increased dTFA with increased BMI and waist circumference fits with other documented adverse metabolic associations of dTFA and is buttressed by experimental documentation that dTFA (without excess calories) increases visceral fat in animals, supporting prospects for causality in the observed association.

## Introduction

Dietary trans fatty acids (**dTFA**), primarily of industrial origin, have been linked to adverse health effects related to lipids, metabolic function, insulin resistance, oxidative stress, inflammation, behavior, and cardiac and general health ^1-15^. Most dTFA are industrially produced via hydrogenation of unsaturated oils, which transform into solids at room temperature ^16^. (Small amounts of naturally occurring dTFA, bearing a different chemical configuration, are ruminant-derived, and may not share the adverse health implications ^17^.)

dTFA has historically been common in many food items, such as margarines, shortenings, baked items and prepared foods, as well as coffee creamers ^18-20^. Because of evidence favoring adverse health implications, coupled with absence of evidence of health benefits, dTFA were first banned in Denmark in 2003 ^21^. FDA calls to label trans fat content in foods were introduced in the US, with labeling required as of January 2006 ^22^. FDA rescinded the designation of dTFA as “generally regarded as safe” in 2015 ^23^. Although final dTFA removal from the US market has a deadline of January 1, 2021 ^24^, dTFA remain in use in many nations throughout the world. Details on which nations currently restrict dTFA, which have plans to do so, and for which nations know such plans are present or known, are available from the World Health Organization (WHO) ^25^. (The WHO has a global initiative entitled Countdown to 2023.) Because dTFA has not been removed from the market globally, effects of dTFA remain a concern in many regions.

It was previously shown that increased consumption frequency (but not amount) of a food that has antioxidant, energy-supporting, and insulin sensitizing effects was linked favorably to body mass index (**BMI**), a finding preserved with adjustment for calories and activity ^26^. This finding in US adults was subsequently replicated in European adolescents ^27^. Conversely, dTFA are prooxidant ^28^, and have been linked to adverse effects on other metabolic factors associated with elevated BMI ^29-32^, factors that favor prospects for an opposite direction effect, and dTFA information was available from the same dataset. In humans, morbidly obese individuals show higher concentrations of trans fatty acids in their visceral fat than do nonobese ^33^. Critically, dTFA administered experimentally in animals (without increased calories) promotes increased visceral fat deposition ^34^.

We sought to evaluate whether higher dTFA consumption was associated with higher BMI, adjusted for calories and activity, capitalizing on cross-sectional data. Because of the animal reports that trans fat feeding increased visceral fat (holding calories constant), we sought in addition to assess the relation of dTFA to waist circumference.

## Methods

### Design

This is a cross-sectional analysis.

### Location

Southern California.

### Participants

Of 1018 adults, 694 were males over age 20, and 324 were surgically or chronologically postmenopausal females, screenees for a clinical trial of lipid lowering therapy ^35 36^. Participants were broadly sampled, however those at the extremes of high or low LDL-cholesterol (>190mg/dL or <115mg/dL), on lipid-lowering medications, with measured fasting glucose >142mg/dL, or with known diabetes, cardiovascular disease, HIV, or cancer were excluded ^35^.

There were no restrictions based on BMI. Mean BMI of participants was similar to that of the general US population at the time of the study. The study protocol was approved by the University of California, San Diego Human Research Protections Program. All participants gave written informed consent. Recruitment, as well as study participation and data collection, occurred from 2000 through 2004, during a privileged period for trans fat assessment, prior to the initiation of trans fat labeling in January 2006.

### Sample Size

This is a secondary analysis of baseline data from a study designed for a different purpose. Original power/sample size/effect size calculations are shown elsewhere ^35^. Assuming 2-sided alpha of 0.05, and 80% power, calculating the sample size for a 1-sample regression, powering for a slope of 0.15 (versus the null hypothesis slope of 0) and setting the standard deviation to 1 (in arbitrary units), the required sample is 343 (G*Power 3.1.9.7 ^37^). This means that our sample size, of 897 for BMI and 901 for waist circumference (combined sex sample) are more than adequate. For men, the sample size for BMI of 603 approaches the size enabling analysis stratified by age (depending on de-facto effect size). This sample size provides 96% power to detect a slope of 0.15 (2-sided alpha 0.05). For women, the sample size of 284 for BMI would suffice only if the effect size is larger than in this calculation. The required slope for women to provide 80% power would be 0.16. Alternatively analyzed, the power would be 72% to detect a slope of 0.15 for our de-facto sample size of 284 (the number of women with dTFA and BMI assessed), under the above parameter specifications.

### BMI calculation

972 (95.6%) participants had both weight and height measured at screening, from which BMI was calculated (kg/m^2^).

### Waist Circumference

971 participants (95.4%) had waist circumference measured at screening (cm). Several waist circumference assessment approaches have been reported to relate similarly to non-waist circumference metabolic syndrome factors and cardiovascular outcomes ^38^. We measured waist at the level of the umbilicus, which has been reported to show the greatest sensitivity in prediction of cardiometabolic risk ^39^.

### Dietary Trans Fatty Acid Estimation

945 participants (92.8%) completed a dietary assessment prior to a baseline visit and are the focus of this analysis. “Nutrient data were collected using a food frequency questionnaire developed by the Nutrition Assessment Shared Resource of the Fred Hutchinson Cancer Research Center” ^40^. Consumption frequency and portion size were elicited, for a suite of food categories, each in turn defined by a set of foods or beverages. Additional questions addressed food preparation and purchasing, to further refine nutrient calculations.

Nutrient calculations were performed using the Nutrient Data System for Research software version 4.03, developed by the Nutrition Coordinating Center, University of Minnesota Food and Nutrient Database (version 31, released November 2000), which added *trans* fatty acid values in 1998 ^1^.

*Trans*-fatty acid values were determined for all foods in the database (0% missing) and include individual contributions of 16:1 *trans* (*trans*-hexadecenoic acid); 18:1 *trans* (*trans*-octadecenoic acid); and 18:2 *trans* (*trans*-octadecadienoic acid), which encompasses *cis-trans, trans-cis*, and *trans-trans* forms; as well as total *trans*-fatty acids. The USDA table “Fat and Fatty Acid Content of Selected Foods Containing Trans-Fatty Acids” – was the primary source of *trans*-fatty acid information for assignment of values to foods in the database ^41^. Additional data sources included other nutrient databases and articles in the scientific literature containing *trans*-fatty acid values for US foods, using appropriate methodologies ^42^.

The study period (1999-2005) was advantageous because dTFA values were available in the nutrient database, while trans fat composition in foods was relatively stable. USFDA trans fat labeling requirements, motivating changes by manufacturers in food composition, were implemented later on January 1, 2006 ^1 22^.

### Covariates

Candidate confounders, of potential relevance to dTFA and to BMI, included age, sex, calorie intake (obtained from the Food Frequency Questionnaire), activity level (measured as times in a week exercised vigorously for at least 20 minutes), chocolate consumption frequency (times eaten per week, which was favorably linked to BMI in this sample ^26^ and elsewhere in the literature ^27^) and mood, assessed by the Center for Epidemiological Studies Depression scale (**CES-D**) (adversely linked to dTFA in this sample ^1^ and in literature ^43^).

### Analyses

Regression analysis with robust standard errors ^44^ was used to assess the relation of dTFA (predictor) to BMI (outcome), unadjusted and in models adjusting for potential confounders, including age and sex, then adding calories and exercise, chocolate frequency, and depression successively. All variables except sex were treated as continuous variables for analysis (not binned). Additional analyses were conducted, stratified by sex; and stratified by age for men. This assesses for effect modification, and in absence of strong effect modification, affords intra-sample replication of dTFA to assessed metabolic outcomes. (Recall women of procreative potential were excluded as these were screenees for a clinical trial, so younger adults were predominantly male.)

Few covariate values were missing, and missing values were not imputed. For both BMI and waist circumference, the fully adjusted model bore 98.9% of the participants from the unadjusted model: 887 out of 897 for BMI; 891 out of 901 for waist circumference. Specifically, among the sample analyzed (those with both dTFA and either BMI or waist circumference assessed), no participants were missing values for age, sex, or calorie intake. 3 were missing values for exercise; with one further lost due to lack of chocolate frequency information, and 6 lost due to missing data for depression (CES-D).

Analyses used Stata version 11.0, College Station, TX. 2-sided p-values <0.05 defined statistical significance.

### Patient and Public Involvement

Patients or the public were not involved in the design, or conduct, or reporting, or dissemination plans of our research.

## Results

Participants were 68.2% male, with age (mean(SD)) 57.0(12.2) years (range 20-85), and BMI 27.9(4.4) kg/m^2^ (range 17.3 to 49.6) (compared to 27.93, the US national average for age greater than 20 with 2/3 men for years 1999-2002) ^45^. Mean dTFA consumption was 3.5(2.5) grams/day (range 0.1 to 27.7). Average times/week of vigorous exercise at least 20 min was 3.6(3.0) (range 0-28). Mean weekly chocolate consumption frequency was 2.0(2.5) times/week (range 0-20). Mean CES-D score was 8.2(7.2) (range 0-52). See **Table 1** for participant characteristics, for the total sample and stratified by sex.

**Table 1.**
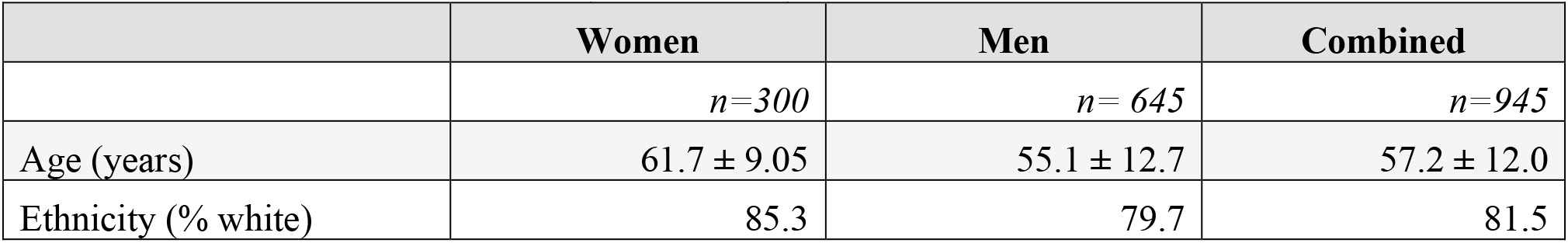
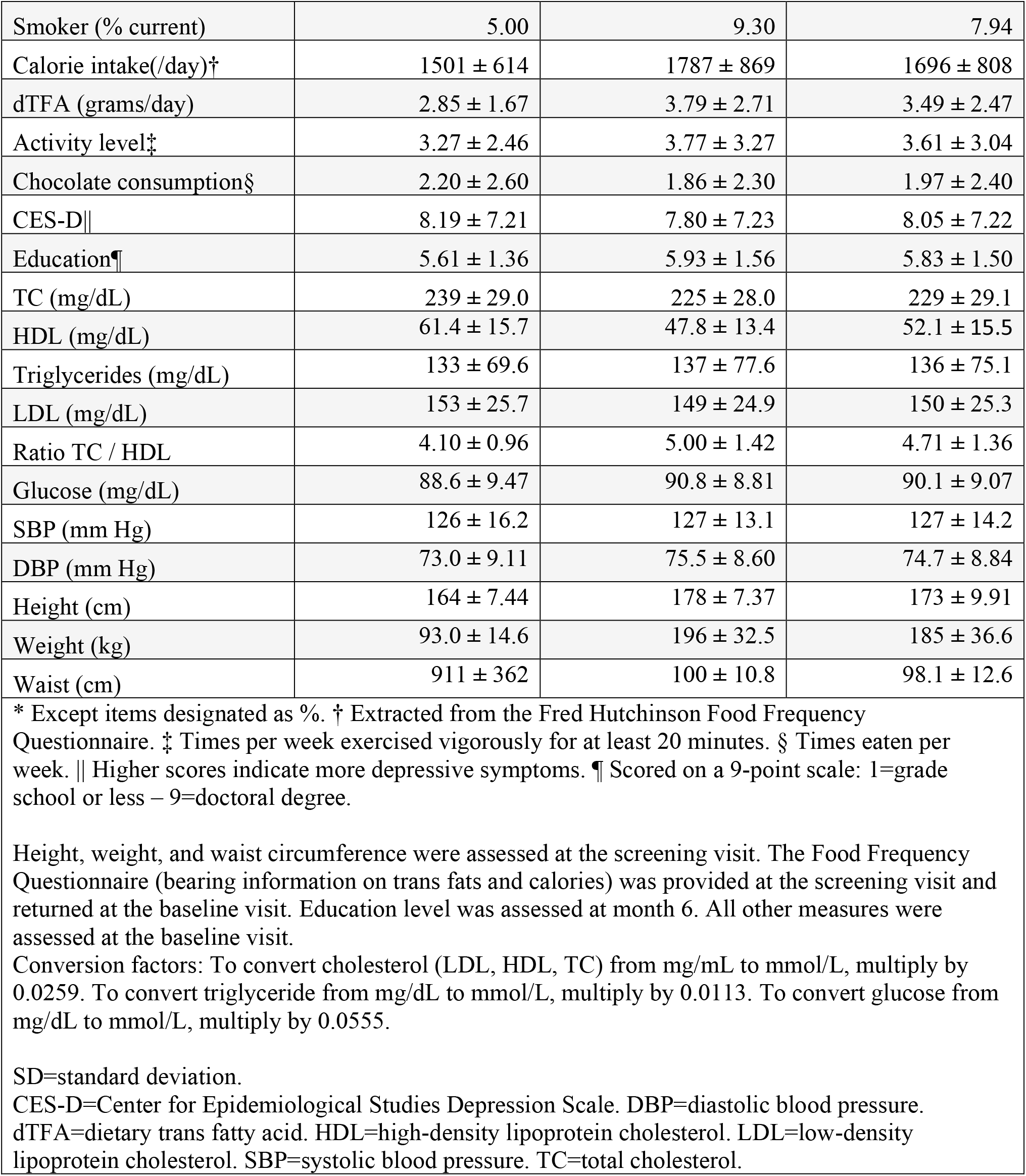
Participant Characteristics (Mean ± SD*)

The relation of trans fat consumption to BMI was positive and strongly significant (**Table 2**). Adjustment for calorie intake and activity did not attenuate, *and indeed strengthened*, the magnitude of the association. In the models adjusted for calorie intake and activity, the magnitude of association was such that each additional 2.5 grams/day of trans fats consumed was associated with an expected one-point higher BMI.

**Table 2.** Trans Fat Consumption Predicts Greater BMI and Waist Circumference: β (Coefficient) is per gram/day dTFA.

|  | BMI (kg/m<sup>2</sup>) |  |  |  | Waist Circumference (cm) |  |  |  |
| --- | --- | --- | --- | --- | --- | --- | --- | --- |
| *Variables Adjusted* | N | $\beta$ (SE) | 95% CI | P | N | $\beta$ (SE) | 95% CI | P |
| Unadjusted | 897 | 0.29 (0.07) | 0.16, 0.42 | **<0.001** | 901 | 0.93 (0.20) | 0.54, 1.3 | **<0.001** |
| Age, sex | 897 | 0.25 (0.07) | 0.11, 0.38 | **<0.001** | 901 | 0.73 (0.20) | 0.34, 1.1 | **<0.001** |
| Age, sex, activity*, calories† | 894 | 0.43 (0.11) | 0.20, 0.65 | **<0.001** | 898 | 1.0 (0.34) | 0.34, 1.7 | **0.003** |
| Age, sex, activity, calories, chocolate‡ | 893 | 0.43 (0.12) | 0.21, 0.66 | **<0.001** | 897 | 1.0 (0.34) | 0.35, 1.7 | **0.003** |
| Age, sex, activity, calories, chocolate, mood (CES-D) | 887 | 0.44 (0.12) | 0.21, 0.67 | **<0.001** | 891 | 1.0 (0.35) | 0.37, 1.7 | **0.003** |
| * Times per week exercised vigorously for at least 20 minutes. † Extracted from the Fred Hutchinson Food Frequency Questionnaire. ‡ Times eaten per week.<br>Regression analysis with BMI (or waist circumference) as the dependent variable, dietary trans fats as focal predictor variable, using robust (aka heteroskedasticity independent, aka White) standard errors<sup>44</sup>.<br>$\beta$=regression coefficient. CI=confidence interval. SE=standard error.<br>BMI=body mass index. CES-D=Center for Epidemiological Studies Depression Scale. dTFA=dietary trans fatty acid. |  |  |  |  |  |  |  |  |

Findings were separately present and significant for men and for women (**Table 3**). Although the magnitude of the coefficient was greater for women, significance was greater for men, who were more numerous. The older typical age of female participants did not appear to explain the stronger coefficient: *Younger* age was associated with appearance of a stronger dTFA-BMI association among men (in whom representation across the adult age spectrum better permitted comparison). Men over 50, age-similar to the women in the sample, had a coefficient approximately half that observed in women.

**Table 3.** Trans Fat Relation to BMI, and to Waist Circumference: Stratified by sex; then stratified by age (among men)

|  | BMI (kg/m<sup>2</sup>) |  |  |  | Waist Circumference (cm) |  |  |  |
| --- | --- | --- | --- | --- | --- | --- | --- | --- |
| *Group Assessed* | **N** | **β (SE)** | **95% CI** | **P** | **N** | **β (SE)** | **95% CI** | **P** |
| Combined sex (as in Table 1) | 887 | 0.44 (0.12) | 0.21, 0.67 | **<0.001** | 891 | 1.0 (0.35) | 0.37, 1.7 | **0.003** |
| Men | 603 | 0.43 (0.13) | 0.18, 0.68 | **0.001** | 606 | 1.1 (0.40) | 0.31, 1.9 | **0.006** |
| Women | 284 | 0.67 (0.29) | 0.10, 1.2 | **0.021** | 285 | 1.3 (0.72) | -0.09, 2.8 | **0.066** |
| Younger Men (age ≤50; n=215) | 215 | 0.56 (0.21) | 0.15, 0.98 | **0.008** | 215 | 1.4 (0.56) | 0.26, 2.5 | **0.016** |
| Older Men (age > 50; n=388) | 388 | 0.34 (0.15) | 0.04, 0.65 | **0.026** | 391 | 0.89 (0.50) | 0.10, 1.9 | 0.08 |
| Mean age for men: 54.8 years (SD 12.8). Mean age for women: 61.7 years (SD 9.2).<br>Mean age for men over 50: 62.8 years (SD 8.2). Mean age for men under 50: 41.4 years (SD 6.3).<br>Regression analysis with BMI as the dependent variable, dietary trans fats as predictor, using robust (aka heteroskedasticity independent, aka White) standard errors<sup>44</sup>.<br>Analyses adjusted for age, sex, activity, calories, chocolate, mood (Center for Epidemiological Studies Depression Scale).<br>β=regression coefficient. CI=confidence interval. SD=standard deviation. SE=standard error.<br>BMI=body mass index. |  |  |  |  |  |  |  |  |

Buttressing the findings for BMI, dTFA also showed a significant positive association to waist circumference, robust again to adjustment model (**Table 3**), paralleling findings for BMI. Again, the coefficients appeared stronger for younger men and for women than for older (i.e. age-similar) men. The magnitude of association was on the order of 1cm greater waist circumference per gram/day greater dTFA, across the total sample (1.4cm for young men).

## Discussion

Greater dTFA was associated with higher BMI in adult men and women. The association was not explained by calorie intake and exercise expenditure – indeed, was strengthened with adjustment for these. Each additional 2.5 grams of trans fats consumed daily was associated with an expected BMI increment of one point. Findings were supported in men and women, and buttressed by parallel associations for waist circumference, in which each additional gram of trans fats consumed daily was associated with an estimated 1cm greater waist circumference (or 1.4cm in men age ≤50). A 2006 report observed it was possible to consume 10-25 grams of dTFA in one day (e.g. consuming fast food fare) ^46^, and have an expected higher BMI by 4-10 points. This is consistent with the largest estimated dTFA, of 28 grams, among participants in our sample.

These findings align with hypotheses dating back ∼ three decades, that dTFA may promote obesity ^47^. They cohere with evidence relating oxidative stressor exposures to elevated BMI ^48^. They comport with evidence from animal studies that visceral fat deposition increases with trans fat intake, in absence of calorie excess ^34^. dTFA was previously linked to waist circumference in men ^31^, and to weight gain in women ^49^ (or in combined sexes, not broken out by sex ^50^). This provides (to our knowledge) the first evidence supporting both effects, and documenting effects in each sex. A common presumption has been that the food contribution to BMI depends exclusively on food calories consumed (or, in reality, *potential* calories available from macronutrients in ingested foods) versus calorie/energy expenditure through activity – perhaps leaving leeway for such factors as muscle and bone mass. By this thinking, dietary changes that are isocaloric cannot influence BMI (holding activity constant). However, substantial evidence illustrates that other exposure factors, beyond caloric input and exercise, influence adiposity ^26 48 51^. Identification of environmental obesogens – like bisphenol A – underscores this ^52 53^. Signaling properties of chemicals in foods can alter assimilation and utilization of calories/energy in protean ways: for example, they may promote diarrhea ^54-59^, alter gut microbiome ^60-62^, affect sleep ^56 63-67^, alter thyroid function ^68^, redirect calories toward or against thermogenesis ^69^, stimulate mitochondrial oxidative phosphorylation capacity ^70-72^, direct utilization of calories to other physiological functions such as blood vessel production ^70^ – with capillarity in turn affecting efficiency of calorie utilization by cells – and/or alter regulation of genes involved in lipolysis or lipogenesis ^73^, moderating the fraction of consumed energy directed to fat deposition.

Findings, in the context of triangulating literature, have implications for international decisions regarding trans fat labeling and restrictions. For nations that follow the former US strategy of labeling any product that has less than 0.5 grams trans fat “per serving” as “0 grams trans fat,” the 2.5 grams/day dTFA associated with one BMI point increment could be achieved with accretion of five “servings” daily of any complement of putatively “0 grams trans fat” foods. (Some food products achieved this by reducing the stated size of a serving ^74^.) Similarly, 2 such “servings” daily would produce the 1gram/day estimated to be associated with 1cm greater waist circumference. We use the term “associated” because observational data are a problematic basis for inferences about causality. It is the triangulation with other evidence, including experimental evidence in animals, that suggests a causal relationship may well be present.

This study is observational; confounding and bias cannot be excluded. Differential recall of diet by obese people is possible – though, the finding would require that recall/reporting bias leads those with higher BMI to report a “worse” quality diet (more trans fats as a fraction of calories) than their lower BMI counterparts, counter to the more expected direction of reporting bias. Alternatively, obese people could differentially crave trans fat laden foods. The study also has strengths, including the sizeable sample, concurrent assessment of weight and waist circumference, segregation of analysis by sex, and the inclusion of key important covariates. A critical advantage is conduct of the study during a privileged time in which trans fat assessment had been added to the food frequency questionnaire, but trans fat consumption was comparatively stable – prior to implementation of FDA mandate for trans fat labeling and public health efforts to curb trans fat intake. Although the study is observational, concordance with findings from experimental animal studies, which are not similarly subject to bias and confounding, adds crucial support to observe associations and their potential causal underpinnings. Observational studies coupled with experimental studies in animals remain vital for examining effects of dTFA (and other exposures recognized or suspected to have adverse implications), as randomized dTFA allocation may be deemed ethically problematic in humans.

The consistency with other observational human studies, but as above more *particularly with experimental findings from nonhuman primates* (in which dTFA without caloric excess increased visceral fat deposition), adds vitally to prospects for causality. With the FDA’s rescission of the designation of dTFA as “generally recognized as safe” ^23^ and the January 2021 closure of the marketing window in the US for trans fat inclusion in foods ^24^, the time window for evaluation even with cross-sectional data is closing (for human studies) in the US. Continued acceptance of dTFA in many nations underscores the continued importance of evidence on health correlates of dTFA. Women of procreative potential, as well as children, were excluded. Though it is possible findings need not apply to these groups, within the sample female sex and younger age were tied to larger apparent magnitude associations of dTFA to BMI, suggesting the possibility that the dTFA-BMI relationship might be not merely present, but conceivably stronger in excluded groups. The cross-sectional character of the study would normally merit a call for prospective validation, and extension to include procreative women and children. If undertaken, such studies must occur in nations beyond the US that have not – or not yet – enacted similar dTFA restrictions.

This study employed dietary recall rather than objective markers of trans fatty acids such as red blood cell membrane trans fats, or plasma phospholipid trans fats ^75 76^. This is a strength as well as a weakness. Biomarkers of nutrients, though objective, may be influenced by factors – known or unknown – distinct from nutrient intake, and such factors may themselves influence health outcomes, which can create the spurious appearance of a connection of the dietary factor to the outcome. For instance, oxidative stressor exposure can alter vitamin D levels at a given level of vitamin D intake ^77^, but oxidative stress also contributes to adverse processes like inflammation ^78 79^, apoptosis ^80 81^ and endothelial dysfunction ^82^ – adversely affecting many outcomes. Use of vitamin D *levels* might thus exaggerate the impact of low vitamin D *itself* on adverse outcomes (and has done so). Or, as previously observed, “low serum levels of iron may be linked to colon cancer death, or death from exsanguination, but it is neither low iron intake nor low iron levels that *produce* these. Rather, colon cancer may cause gastrointestinal bleed and low measured iron *at a given iron intake level*, producing the appearance of a connection” ^1^. These are illustrations of a general principle: Processes that alter objective marker levels of a dietary constituent, independent of intake, can also influence outcomes, which can lead to spurious appearance (or disappearance) of a connection between dietary intake of the compound and the outcome. Thus, use of such a measure comes with peril.

dTFA was estimated from dietary recall, and foods of the same name need not reliably have had the same trans fat content. However, provided any misclassification is nondifferential, this would be expected to produce bias toward the null; that is, the “true” relationship of dTFA to BMI might be stronger still. Additionally, data were collected during a period of relative stability of trans fat content in foods (2000-2004), prior to subsequent efforts to label (2006) and restrict dTFA – a relative strength for assessing dTFA associations ^1^.

Findings from this study, linking dTFA to higher BMI (as well as waist circumference), add to an extensive body of evidence supporting adverse health associations of dTFA. The premise of the study rests on a robust biological foundation. Factors including the strength of association, biological gradient, biological plausibility, and coherence with other literature add (ahem) “weight” to the possibility that the association we identify may have a causal basis.

## Supporting information

Strobe Checklist

## Data Availability

Statistical code and print out available from corresponding author.

## Acknowledgements

We gratefully acknowledge the assistance of all the UC San Diego Statin Study personnel, particularly Janis Ritchie BSN and Julie Denenberg MA; as well as the staff at the UC San Diego General Clinical Research Center. We also thank our participants for generously giving their time. All authors had full access to all the data in the study and take responsibility for the integrity of the data and the accuracy of the data analysis.

## Author Contributions

Conceived and designed the study, BG; Analyzed the Data, BG; Replicated/Validated the Analyses, HK; Wrote the Paper, BG; Critical Revisions of Manuscript, HK, AP; Layouts, HK, AP; Updating Discussion/Citations: AP; Final approval, BG, HK, AP; Funding Acquisition, BG

## Original study protocol

Not applicable.

## Data availability statement

Statistical code and print out available from corresponding author.

## Patient consent for publication

Not applicable.

## Provenance and peer review

Not commissioned; externally peer reviewer.

